# Estimation of the real magnitude of antimicrobial resistance of *Mycoplasma genitalium* in Belgium by implementing a prospective surveillance programme

**DOI:** 10.1101/2023.06.06.23291033

**Authors:** Irith De Baetselier, Hilde Smet, Kaat Kehoe, Imelda Loosen, Marijke Reynders, Iqbal Mansoor, Lorenzo Filippin, Mathieu Cauchie, Ellen Van Even, Nadia Makki, Gilberte Schiettekatte, Wouter Vandewal, Bart Glibert, Veerle Matheeussen, Yolien Van der Beken, Reinoud Cartuyvels, Sophia Steyaert, Ann Lemmens, Maria-Grazia Garrino, Henry Paridaens, Elena Lazarova, Bénédicte Lissoir, Marine Deffontaine, Amélie Heinrichs, Veroniek Saegeman, Elizaveta Padalko, Amaryl Lecompte, Wim Vanden Berghe, Chris Kenyon, Dorien Van den Bossche

**Author notes:** Corresponding author: Dr. Irith De Baetselier, Institute of Tropical Medicine, National Reference Centre of Sexually transmitted infections Belgium, Department of Clinical Sciences, Nationalestraat 155, 2000 Antwerp, Belgium.

## Abstract

**Objectives:** Antimicrobial resistance of *Mycoplasma genitalium* (MG) is a growing concern worldwide. Because reliable data on the burden of resistant MG in Belgium are missing, an additional prospective surveillance program was implemented in 2022 to estimate the real burden of resistant MG in Belgium.

**Methods:** Belgian laboratories (n=21) provided frozen remnants of MG positive samples to the National Reference Centre of Sexually Transmitted Infections from July to November 2022. The presence of macrolide and fluoroquinolones resistance associated mutations (RAMs) was assessed using Sanger sequencing of the 23SrRNA and *parC* gene. Differences in resistance patterns were correlated with surveillance methodology, socio-demographic and behavioral variables via Fisher’s exact test and logistic regression analysis.

**Results:** Sequencing for both macrolide and fluoroquinolone RAMs was successful for 232/244 MG positive samples. Over half of the samples were resistant to macrolides (55.2%). All MG in samples from men who have sex with men (MSM) (24/24) were resistant to macrolides. The presence of fluoroquinolone RAMs was estimated to be 26% and did not differ with socio-demographic and sexual behaviour characteristics.

**Conclusions:** Given the considerable cost of macrolide resistance testing, our data suggest that the use of macrolide resistance testing in MSM does not seem justified in Belgium. However, the lower prevalence of macrolide resistance in other population groups, combined with further emergence of fluoroquinolone resistance provides evidence for macrolide resistance testing in these groups. Continued surveillance of resistance in MG in all groups will be crucial to guide national testing- and treatment strategies.

## Introduction

*Mycoplasma genitalium* (MG) is a sexually transmitted infection (STI). In high-resource countries the prevalence of MG is estimated to be 1.3% in the general population without significant differences between men and women.[1] In Belgium, previous studies have reported higher prevalence figures but these were estimated only among populations who are behaviourally more at risk such as men who have sex with men (MSM) (17.2%) and female sex workers (10.8%).[2,3] In a recent Dutch study, a prevalence of 13.8% was found among individuals visiting a STI clinic and it ranged from 8.2% among men who have sex with women (MSW), 12.6% in women to 20.1% among MSM which is higher than previously reported.[4]

Although MG is estimated to be prevalent in sexually active individuals, the majority of infections are asymptomatic and can clear spontaneously. Consequently, data on sequelae from untreated infections remain too limited to implement MG screening.[5,6] Moreover, of concern is the ability of MG to acquire resistance to azithromycin and moxifloxacin, which are currently the most prescribed antimicrobials to treat MG infections.[7] Doxycycline, the recommended first-line therapy for non-gonococcal urethritis in Europe and the US, has a low cure rate of 30-40% and is now predominantly used in two-stage therapy approaches for MG.[6,8,9] These approaches use doxycycline first to lower the bacterial load of MG and afterwards, a second antibiotic (azithromycin or moxifloxacin) is administered according to the pathogen’s susceptibility profile.[5]

In Belgium, we previously reported a high macrolide resistance rate of 68.3% in MG. In addition, 18.0% of the samples also harboured resistance associated mutations (RAMs) to fluoroquinolones.[10] However, these numbers may not accurately reflect the resistance rates in the general population since most samples were from MSM (95/167; 56.9%).

Since 2015, the National Reference Centre of Sexually Transmitted Infections (NRC-STI) in Belgium has included the confirmation of MG as one of its surveillance tasks. However, since 2018, a strict testing strategy has been applied in order to follow the testing guidelines of the International Union against STIs (IUSTI). This testing strategy entails that testing is performed, in most cases, only in the event of persistent urethritis/cervicitis and when chlamydia/gonorrhoea has been excluded.[7] As a consequence, the yearly number of samples tested is low and may not be representative of the entire Belgian population.

In France, a systematic prospective collection of MG positive samples of approximately 20 public and private microbiology diagnostic laboratories is performed yearly since 2018 to estimate the magnitude of MG antimicrobial resistance (AMR).[11]. This surveillance method may provide a more comprehensive picture of the degree of MG AMR. The presence of macrolide RAMs was estimated to be 60% in men and 22% in women in 2020 in France. Fluoroquinolone RAMs were present in 17% and in 15% of the male and female samples respectively, which is lower than the estimates in Belgium.[11]

The aim of this study was to implement a similar surveillance method in Belgium to estimate the exact magnitude of MG antimicrobial resistance. Moreover, we explored the current surveillance method’s accuracy in assessing the burden of MG in Belgium.

## Methods

### National surveillance of MG in Belgium

In Belgium, laboratories can send suspected MG positive samples for analysis to the NRC-STI only when the request is in accordance with IUSTI 2021 guidelines.[7]

DNA extraction is performed using the Abbott m2000sp instrument (Abbott Molecular, Des Plaines, Illinois, USA) and CT/NG extraction kit according to manufacturers’ instructions. MG detection is performed using the S-DIaMGTV multiplex kit (Diagenode Diagnostics, Seraing, Belgium) using the Abbott m2000rt platform.

### Molecular detection of macrolide and fluoroquinolone resistance associated mutations

Specific in-house PCRs were used to retrieve amplicons of 23SrRNA and the parC gene using the GeneAmp PCR System 2700 (Applied Biosystems).[12,13] DNA extraction was performed using the same methodology as mentioned above. Sanger sequencing of the 23S rRNA and parC genes to detect the presence of RAMs to macrolides and fluoroquinolones respectively was performed using the ABI 36730xl instrument of Applied Biosystems (USA) as previously described at The Neuromics Support Facility – VIB, Antwerp, Belgium.[10] Sequences were aligned and analysed by two independent readers using BioEdit Sequence Alignment Editor (Nucleics.com). The following alterations in ParC were coded to be resistant to fluoroquinolones (S83I, S83R, D87N, D87Y).

### Prospective MG-AMR study design

All Belgian laboratories were contacted and asked to complete a questionnaire concerning MG testing and their willingness to participate in the prospective study. The survey included questions concerning the molecular testing technique, restriction rules applied to MG testing if applicable, and the number of samples tested between 01 January and 31 March 2022 (Q1) including positivity ratio. Participating laboratories were asked to send approximately 12 frozen remnants of consecutively collected MG positive samples to the NRC-STI during July-November 2022. An additional questionnaire with socio-demographic and sexual behaviour characteristics was completed for each positive sample. Samples were tested at the NRC-STI for the presence of macrolide and fluoroquinolone RAMs as mentioned above.

Laboratories analysing over 1000 samples in Q1 were asked to provide positivity ratios for 2022 as well as information regarding their testing strategy.

The prospective study has been approved by the Institutional review board of the Institute of Tropical Medicine (ITM) (ref: 1594/22) and the Ethics Committee of the University of Antwerp (ref: 3534).

### Statistical methods

A descriptive analysis was performed for all MG cases. In case of returning visits, only results with an interval of > 90 days were included in the analysis. Resistance patterns (macrolides, fluoroquinolones or both) were correlated with surveillance method, geographical location, gender, transmission and the presence of symptoms through Fisher’s exact test. If statistical significance was detected (p<0.05), this variable was included in a multivariate logistic regression model. Adjusted odds ratio’s (aOR) with their corresponding 95% Confidence Intervals (CI) were reported. STATA V15.1 (StataCorp LP, College Station, Texas, USA) was used for all statistical analyses. A significance level of 0.05 was used.

## Results

### MG testing strategy in Belgium

A total of 21 hospital- and private laboratories participated in the study and each region of Belgium was represented (Supplementary Figure 1). Supplementary Table 1 documents more details of the laboratories, including the number of samples tested during Q1 2022, the number of positive samples, the molecular technique of MG detection and the number of samples shipped to the NRC-STI for further AMR testing. MG testing was mostly performed in the framework of a broader STI screening by performing a multiplex assay which includes at minimum *Chlamydia trachomatis, Neisseria gonorrhoeae, Mycoplasma genitalium* and *Trichomonas vaginalis* (16/21 laboratories) or at the clinician’s request. Positivity ratio of MG in Q1 2022 varied between 0 and 14.0%.

### MG prevalence estimates in Belgium

The NRC-STI received 351 samples for MG testing during the year 2022. A total of 29.1% (102/351) of the requests did not comply with the testing guidelines and were discarded. Of the remaining 249 samples, 36 were found to be positive for MG leading to a positivity ratio of 14.5%. The MG positivity ratio’s, stratified by gender, obtained by the NRC-STI and by the four laboratories that tested over 1000 samples in the first quarter of 2022 are tabulated in Table 1.

**Table 1:** Prevalence rates of M. genitalium in 2022 stratified between gender in different laboratories in Belgium

| Laboratory | Location | All | Male samples | Female samples | Testing strategy |
| --- | --- | --- | --- | --- | --- |
| NRC-STI | Flanders | 36/249<br>(14.5%) | 27/180<br>(15.0%) | 9/69<br>(13.0%) | Only samples of symptomatic individuals where CT/NG is excluded |
| Private Laboratory | Flanders | 287/35 927<br>(0.80%) | 91/12 265<br>(0.70%) | 196/23 466<br>(0.80%) | All samples for CT/NG testing or MG testing. Results will only be provided when CT/NG is negative |
| Private Laboratory | Wallonia | 121/2527<br>(4.8%) | 16/260<br>(6.1%) | 105/2267<br>(4.6%) | All samples for CT/NG testing or MG testing. |
| Hospital Laboratory | Flanders | 61/1214<br>(5.0%) | 43/313<br>(13.7%) | 18/901<br>(2.0%) | All samples of symptomatic individuals (STI clinics), STI screening in risk populations (including young adults <25 years), together with pregnancy screening in risk populations |
| Private Laboratory | Wallonia | 401/8385<br>(4.8%) | unknown | unknown | All samples for CT/NG testing or MG testing. |

NRC-STI: National Reference Center for Sexually Transmitted Infections; CT/NG: *Chlamydia* *trachomatis*/*Neisseria gonorrhoeae*; MG: *Mycoplasma genitalium*

### Demographics and behavioural characteristics of MG positive samples

Besides the 36 samples detected by the NRC-STI, 208 samples were collected during the prospective study, providing a total number of 244 positive samples. The socio-demographic, behavioural characteristics and symptomatology pertaining to the samples can be found in Table 2. Almost all samples were of urogenital origin (229/244; 93.9%) and more than half were obtained from women (154/244; 63.1%). Most of the samples were collected by a gynaecologist (90/208; 43.3%), followed by a general practitioner (75/208; 36.1%) and 18 samples were taken by other clinicians such as urologists, infectiologists, dermatologists, surgeons or emergency doctors. Of the cases with reported symptomatology, 66.5% (123/185) represented urogenital symptoms, but 25.9% (48/185) were from asymptomatic individuals. Notably, MG was also present in patients with vague symptoms such as vaginal pain, blood loss, burning sensation, irritation, abdominal pain and discharge (31.4%; 58/185).

**Table 2:**
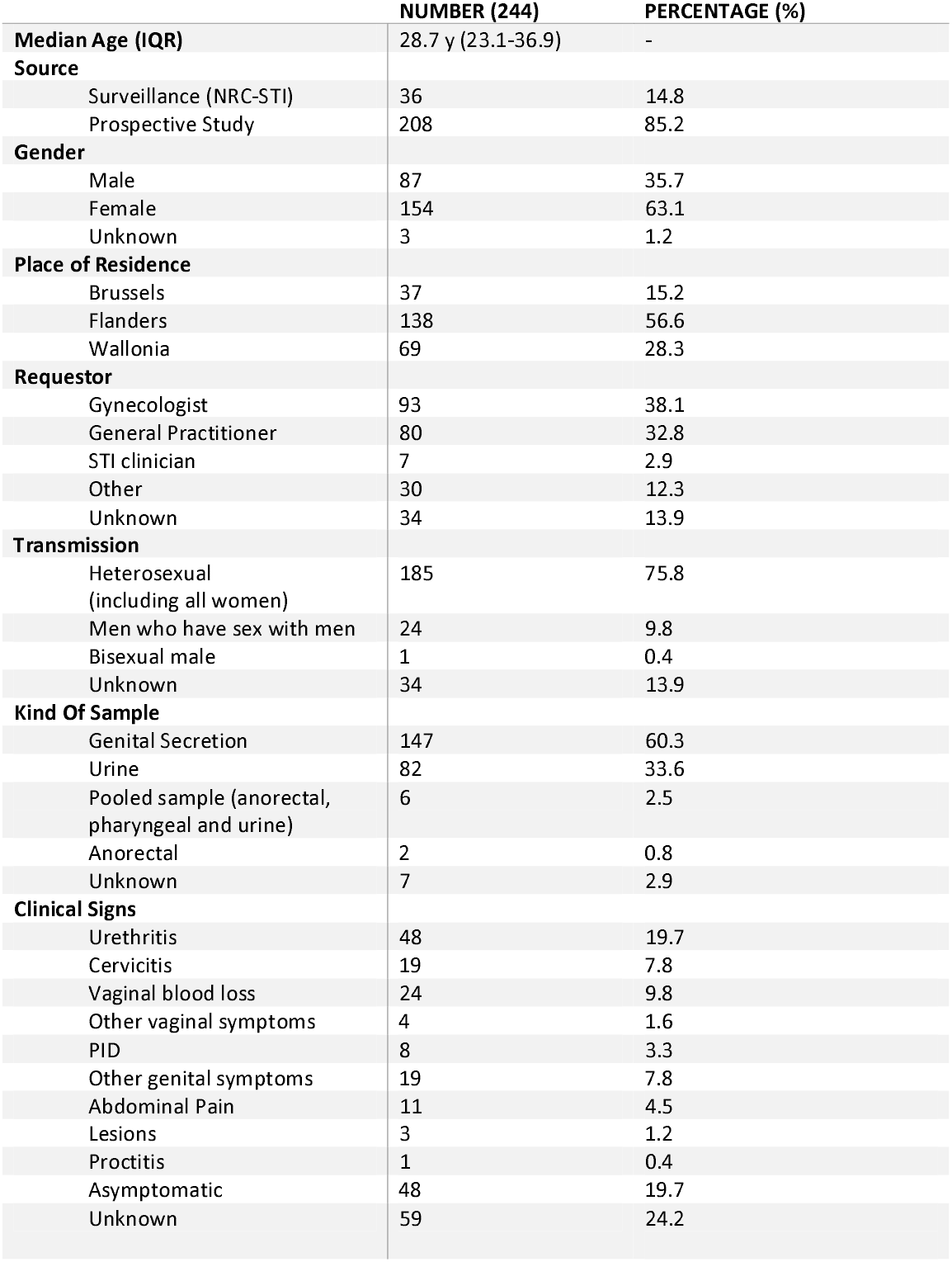
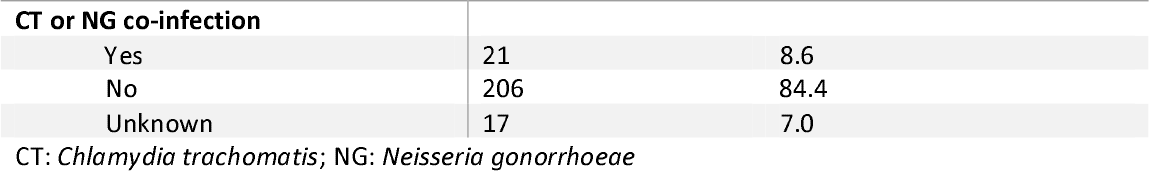
Socio-demographic and behavioral characteristics of M. genitalium positive samples

|  | NUMBER (244) | PERCENTAGE (%) |
| --- | --- | --- |
| **Median Age (IQR)** | 28.7 y (23.1-36.9) | - |
| **Source** |  |  |
| Surveillance (NRC-STI) | 36 | 14.8 |
| Prospective Study | 208 | 85.2 |
| **Gender** |  |  |
| Male | 87 | 35.7 |
| Female | 154 | 63.1 |
| Unknown | 3 | 1.2 |
| **Place of Residence** |  |  |
| Brussels | 37 | 15.2 |
| Flanders | 138 | 56.6 |
| Wallonia | 69 | 28.3 |
| **Requestor** |  |  |
| Gynecologist | 93 | 38.1 |
| General Practitioner | 80 | 32.8 |
| STI clinician | 7 | 2.9 |
| Other | 30 | 12.3 |
| Unknown | 34 | 13.9 |
| **Transmission** |  |  |
| Heterosexual<br>(including all women) | 185 | 75.8 |
| Men who have sex with men | 24 | 9.8 |
| Bisexual male | 1 | 0.4 |
| Unknown | 34 | 13.9 |
| **Kind Of Sample** |  |  |
| Genital Secretion | 147 | 60.3 |
| Urine | 82 | 33.6 |
| Pooled sample (anorectal,<br>pharyngeal and urine) | 6 | 2.5 |
| Anorectal | 2 | 0.8 |
| Unknown | 7 | 2.9 |
| **Clinical Signs** |  |  |
| Urethritis | 48 | 19.7 |
| Cervicitis | 19 | 7.8 |
| Vaginal blood loss | 24 | 9.8 |
| Other vaginal symptoms | 4 | 1.6 |
| PID | 8 | 3.3 |
| Other genital symptoms | 19 | 7.8 |
| Abdominal Pain | 11 | 4.5 |
| Lesions | 3 | 1.2 |
| Proctitis | 1 | 0.4 |
| Asymptomatic | 48 | 19.7 |
| Unknown | 59 | 24.2 |
| **CT or NG co-infection** |  |  |
| Yes | 21 | 8.6 |
| No | 206 | 84.4 |
| Unknown | 17 | 7.0 |

CT: *Chlamydia trachomatis*; NG: *Neisseria gonorrhoeae*

### Antimicrobial resistance in MG

Sequencing for both macrolide and fluoroquinolone RAMs was successful for 232/244 (95.1%) samples, of which 34 samples (14.7%) were collected in the framework of the NRC-STI MG surveillance and 198 (85.3%) in the prospective study. Figure 1 depicts the antimicrobial resistance patterns of all samples stratified by collection method (NRC-STI or prospective study) and Table 3 shows the antimicrobial resistance patterns for the examined characteristics.

**Figure 1:**
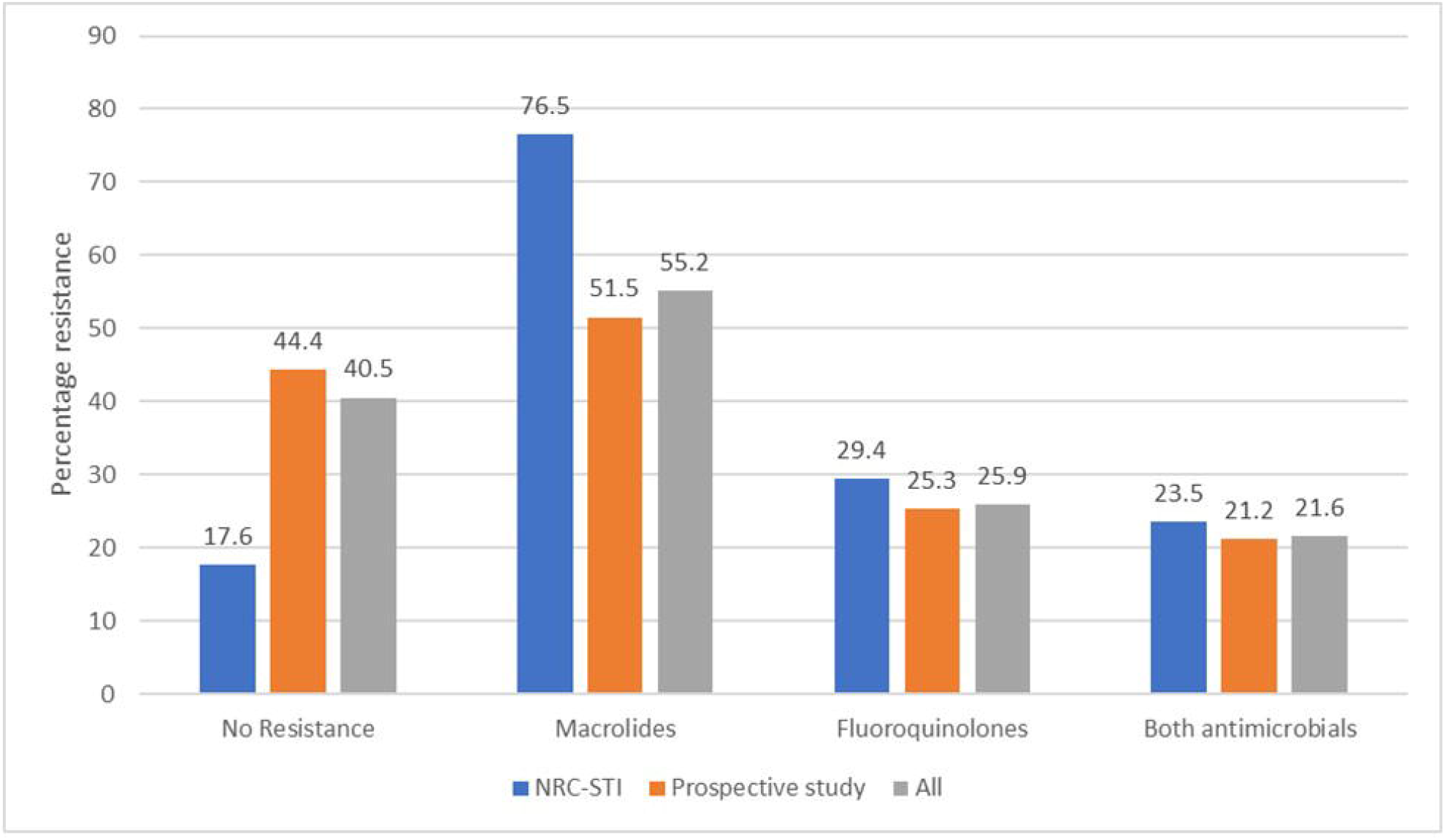
Prevalence of antimicrobial resistance in M. genitalium estimated using samples received by the National Reference Center of STIs (NRC-STI) or in the prospective study or both.

**Table 3:** Socio-demographic characteristics and antimicrobial patterns of M. genitalium

|  | Total |  | Wild type |  | p-value | Macrolide resistant<br>(23S rRNA) |  | p-value | Fluoroquinolone resistant<br>(ParC pos. 83/87) |  | p-value | Multi-resistant<br>(23S rRNA + ParC pos. 83/87) |  | p-value |
| --- | --- | --- | --- | --- | --- | --- | --- | --- | --- | --- | --- | --- | --- | --- |
|  | No. | % | N | % (95% CI) |  | No. | % (95% CI) |  | No. | % (95% CI) |  | No | % (95% CI) |  |
| **All samples** | **232** | **-** | **94** | **40.5(34.1-47.1)** |  | **128** | **55.2 (48.5-61.7)** |  | **60** | **25.9 (20.4-32.0)** |  | **50** | **21.6 (16.4-27.4)** |  |
| **Origin of samples** |  |  |  |  |  |  |  |  |  |  |  |  |  |  |
| NRC-STI | 34 | 14.7 | 6 | 17.6 (6.8-34.5) |  | 26 | 76.5 (58.8-89.3) |  | 10 | 29.4 (15.1-47.5) |  | 8 | 23.5 (10.7-41.2) |  |
| Prospective study | 198 | 85.3 | 88 | 44.4(37.4-51.7) | **0.004** | 102 | 51.5 (44.3-58.7) | **0.008** | 50 | 25.3 (19.4-31.9) | 0.672 | 42 | 21.2 (15.7-27.6) | 0.822 |
| **Place of residence** |  |  |  |  |  |  |  |  |  |  |  |  |  |  |
| Brussels | 35 | 15.1 | 6 | 17.1 (6.6-33.6) |  | 27 | 77.1 (59.9-89.6) |  | 12 | 34.3 (19.1-52.2) |  | 10 | 28.6 (14.6-46.3) |  |
| Flanders | 130 | 56.0 | 55 | 42.3 (33.7-51.3) |  | 71 | 54.6 (45.7-63.4) |  | 36 | 27.7 (20.2-36.2) |  | 32 | 24.6 (17.5-32.9) |  |
| Wallonia | 67 | 28.9 | 33 | 49.3 (36.8-61.8) | **0.005** | 30 | 44.8 (32.6-57.4) | **0.007** | 12 | 17.9 (9.6-29.2) | 0.146 | 8 | 11.9 (5.3 – 22.2) | 0.055 |
| **Gender** |  |  |  |  |  |  |  |  |  |  |  |  |  |  |
| Women | 143 | 61.6 | 71 | 49.7 (41.2-58.1) |  | 64 | 44.8 (36.4-53.3) |  | 36 | 25.2 (18.3-33.1) |  | 28 | 19.6 (13.4-27.0) |  |
| Men* | 87 | 37.5 | 22 | 25.3 (16.6-35.7) |  | 63 | 72.4 (61.8-81.5) |  | 23 | 26.4 (17.6-37.0) |  | 21 | 24.1 (15.6-34.5) |  |
| Unknown | 2 | 0.9 | 1 | 0.5 (1.3-98.7) | **<0.001** | 1 | 0.5 (1.3-98.7) | **<0.001** | 1 | 0.5 (1.3-98.7) | 0.598 | 1 | 0.5 (1.3-98.7) | 0.270 |
| **Transmission** |  |  |  |  |  |  |  |  |  |  |  |  |  |  |
| Women\$ | 143 | 61.6 | 71 | 49.7 (41.2-58.1) | | 64 | 44.8 (36.4-53.3) | | 36 | 25.2 (18.3-33.1) | | 28 | 19.6 (13.4-27.0) | |
| MSM* | 24 | 10.3 | 0 | 0 (0-14.2) |  | 24 | 100 (85.8-100.0) |  | 6 | 25.0 (9.8-46.7) |  | 6 | 25.0 (9.8-46.7) |  |
| MSW | 30 | 12.9 | 11 | 36.7 (19.9-56.1) |  | 18 | 60.0 (40.6-77.3) |  | 10 | 33.3 (17.3-52.8) |  | 9 | 30.0 (14.7-49.4) |  |
| MSU or<br>unknown gender | 35 | 15.1 | 12 | 34.3 (19.1-52.2) | **<0.001** | 22 | 62.9 (44.9-78.5) | **<0.001** | 8 | 22.9 (10.4-40.1) | 0.776 | 7 | 20.0 (8.4-36.9) | 0.574 |
| **STI associated symptoms** |  |  |  |  |  |  |  |  |  |  |  |  |  |  |

|  | Total |  | Wild type |  |  | Macrolide resistant |  |  | Fluoroquinolone resistant |  |  | Multi-resistant |  |  |
| --- | --- | --- | --- | --- | --- | --- | --- | --- | --- | --- | --- | --- | --- | --- |
|  | No. | % | N | % (95% CI) | p-value | No. | % (95% CI) | p-value | No. | % (95% CI) | p-value | No | % (95% CI) | p-value |
|  |  |  |  |  |  | (23S rRNA) |  |  | (ParC pos. 83/87) |  |  | (23S rRNA + ParC pos. 83/87) |  |  |
| No | 45 | 19.4 | 21 | 46.7 (31.7-62.1) |  | 21 | 46.7 (31.7-62.1) |  | 12 | 26.7 (14.6-41.9) |  | 9 | 20.0 (9.6-34.6) |  |
| Yes | 133 | 57.3 | 52 | 39.1 (30.8-47.9) |  | 76 | 57.1 (48.3-65.7) |  | 34 | 25.6 (18.4-33.8) |  | 29 | 21.8 (15.1-29.8) |  |
| Unknown | 54 | 23.3 | 21 | 38.9 (25.9-53.1) | 0.657 | 31 | 57.4 (43.2-70.8) | 0.440 | 14 | 25.9 (15.0-39.7) | 1.000 | 12 | 22.2 (12.0-35.6) | 0.976 |
| **CT or NG infection** |  |  |  |  |  |  |  |  |  |  |  |  |  |  |
| No | 196 |  | 82 | 41.8 (34.8-49.1) |  | 106 | 54.1 (46.8-61.2) |  | 49 | 25.0 (19.1-31.7) |  | 41 | 20.9 (15.4-27.3) |  |
| Yes | 20 |  | 9 | 45.0 (23.1-68.5) |  | 11 | 55.0 (31.5-76.9) |  | 4 | 20.0 (5.7-43.7) |  | 4 | 20.0 (5.7-43.7) |  |
| Unknown | 16 |  | 3 | 18.8 (4.0-45.6) | 0.815 | 11 | 68.8 (41.3-89.0) | 1.000 | 7 | 43.8 (19.8-70.1) | 0.788 | 5 | 31.3 (11.0-58.7) | 1.000 |

\*: including one transwoman. \$: including one woman who reported lesbian transmission. MSM: Men who have sex with men; MSW: Men who have sex with women; MSU: Men with unknown sex of sex partners or unknown gender

Macrolide resistance was found in 55.2% of all samples, however, macrolide resistance was higher in samples collected in the framework of the NRC-STI surveillance; it was the highest in men and in Brussels. All MG from samples collected from MSM harboured RAMs associated with macrolide resistance. Macrolide resistance was also higher in MSW (60.0%) than in women (44.8%).

There was little variation in fluoroquinolone resistance between demographic groups. MSW presented with the highest proportion of fluoroquinolone RAMs (30.0%) compared to MSM (25.0%), but this difference was not found to be statistically significant. In total, more than one in four of the samples had RAMs to fluoroquinolones and one in five had combined fluoroquinolone and macrolide RAMs. Besides the clear association between macrolide resistance and MSM, the multivariate logistic regression model showed that macrolide resistance was less prevalent among women (aOR: 0.36; 95%CI: 0.20-0.64).

Table 4 tabulates the RAMs found in the study stratified per sexual transmission. Interestingly, A2058G was more frequently found among women and MSW whereas A2059G was the most dominant mutation among MSM and men with unknown sex of partners or unknown gender (MSU). D87N and S83I were the most frequently found alterations in ParC. The following combinations of RAMs to both antimicrobials were the most frequently found: A2058G/D87N (48.0%; 24/50) and A2059G/S83I (36.0%; 18/50).

**Table 4:** Resistance associated mutations to macrolides (23S rRNA) and fluoroquinolones (alterations in ParC)

|  | Transmission |  |  |  |  |
| --- | --- | --- | --- | --- | --- |
| Mutation | Women | MSM | MSW | MSU | Total |
| 23S rRNA (*E. coli* numbering) |  |  |  |  |  |
| A2058C | 0 (0) | 0 (0) | 0 (0) | 1 (4.5) | 1 (0.8) |
| A2058G | 32 (50.0) | 5 (20.8) | 8 (44.4) | 4 (18.2) | 49 (38.3) |
| A2058T | 13 (20.3) | 4 (16.7) | 3 (16.7) | 3* (13.6) | 23 (18.0) |
| A2059C | 1 (1.6) | 0 (0) | 0 (0) | 0 (0) | 1 (0.8) |
| A2059G | 18 (28.1) | 15 (62.5) | 7 (38.9) | 14 (63.6) | 54 (42.2) |
| Alterations in ParC |  |  |  |  |  |
| D87N | 21 (56.8) | 3 (42.9) | 5 (50.0) | 0 (0) | 29 (46.8) |
| D87Y | 3 (8.1) | 0 (0) | 0 | 0(0) | 3 (4.8) |
| S83I | 12 (32.4) | 3 (42.9) | 5 (50.0) | 7 (87.5) | 27 (43.6) |
| S83R | 0 (0) | 0 (0) | 0 (0) | 1 (12.5) | 1 (1.6) |
| S83C\$ | 0 (0) | 1 (14.3) | 0 (0) | 0 (0) | 1 (1.6) |
| S83N\$ | 1 (2.7) | 0 (0) | 0 (0) | 0 (0) | 1 (1.6) |

\*mixed infection of A2058G & A2058T; \$ these mutations were not accounted as possible
fluoroquinolone resistant. MSM: Men who have sex with men. MSW: Men who have sex with women. MSU: Men with unknown sex of sex partners or unknown gender.

## Discussion

Our results demonstrate that macrolide resistant MG is rapidly spreading to different populations in Belgium. Half of the samples were resistant to macrolides (55.2%). Although limited in sample size, we report here that 100% of the MG found in samples collected from MSM harboured RAMs to macrolides which is probably related with the use of macrolides to treat other STIs or other infections.[14] These treatment regimens (i.e. one dose of 1g azithromycin) are sub-optimal for MG and may promote the development of AMR and success of macrolide resistant clones.[7] Last year, treatment regimens for STIs were adapted in many countries, including Belgium, to avoid the usage of azithromycin and subsequently, to avoid the further emergence of azithromycin resistance in MG but also in other STIs.[15,16]

Macrolide resistance in MG is now reaching 45% among women. Importantly, our study also identified differences in the distribution of RAMs among different populations. For example, RAM A2059G in 23SrRNA was the most dominant RAM found among MSM, while A2058G was most frequently found among women and MSW. These differences suggest that the circulation of different genotypes of MG may contribute to the spread of macrolide resistance in specific populations. Indeed, a previous phylogenetic analysis in Spain revealed that the spread of strains harbouring a A2058T mutation contributed to the increase in macrolide resistance.[17] Comparing our data with previously published data of 2018, we note an increase in the proportion of samples harbouring this A2058T mutation from 5.0% to 18.0%.[10] This increase was also noted in the Netherlands.[18] This finding highlights the need for additional genetic characterisation of MG to better understand the circulation of the genetic profiles of MG and possible clonal spread of macrolide resistant MG in the different populations.

With the increasing availability of multiplex testing to screen for STIs in Belgium, new recommendations for MG testing and treatment are urgently needed. The prevalence of MG is dependent on the testing strategy used. At the NRC-STI, where only samples of symptomatic individuals are tested, MG positivity rate was nearly 15.0%. However, lower positivity rates were found among laboratories that test MG in all samples collected for STI screening (0.8 to 5.0 %). These estimates may reflect the actual prevalence of MG in the general population and are consistent with rates found in other European countries.[19,20]

The utilization of multiplex technologies that include MG in STI screening panels may result in an excessive use of antimicrobials, thereby promoting resistance.[21] Consequently, it may be recommended to limit the use of such assays. Nevertheless, several diagnostic companies are developing CT/NG/MG/TV multiplex assays. To address this issue, a potential solution is to report the result of MG only upon specific request by the clinician. Furthermore, it is essential to train gynaecologists and general practitioners to test only when necessary, considering that our study found that these specialists were the primary requesters of MG testing.

Importantly, our data may alter testing- and treatment guidelines. We here show that the use of macrolide resistance testing may be of no practical value among MSM in Belgium. However, macrolide resistance testing is still recommended in other populations.

In addition to macrolide resistance, we also observed an increasing prevalence of fluoroquinolone resistance among MG in Belgium. Specifically, we found that 26% of the MG strains had RAMs to fluoroquinolones, with women showing a nearly three-fold increase in fluoroquinolone resistance in MG since 2018 (from 9.1 to 25.2%). Although limited sample size, we also observed that 33% of the MG isolated from samples of MSW harboured fluoroquinolone RAMs, but 40% of these strains were still susceptible to azithromycin. These data indicate that resistance-guided therapy should be implemented in specific populations to ascertain that the appropriate antibiotic is selected.

Unfortunately, in Belgium, sexual behaviour data are mostly not available to the laboratory in order to decide whether or not resistance guided testing is needed. Commercially available AMR assays exist and allow a more rapid turnaround time compared to sequencing, though these are expensive and mostly limited to detection of macrolide RAMs. Moreover, there is currently no reimbursement for MG testing nor for resistance guided testing which in turn may become expensive for the patients. The NRC-STI is performing these tests as part of their reference tasks but with very strict testing rules.

Our results reinforce the need for revision of the MG testing guidelines and reimbursement rules aiming at MG testing only in case of persisting symptoms according to IUSTI guidelines. Macrolide resistance testing in MG positive samples should be implemented for patient groups other than MSM to limit the use of fluoroquinolones to avoid emergence of multidrug resistant MG. One-fifth of our samples presented with RAMs against both antimicrobials. Because the presence of fluoroquinolone RAMs is not always associated with therapeutic failure, some patients can still successfully be treated with moxifloxacin. [22] However, clinical multidrug resistant MG cases are increasingly being reported, also in Belgium.[20, unpublished data] Third-line therapies include minocycline, pristinamycin and even chloramphenicol.[7,24] However, the latter two are difficult to procure in Belgium and novel treatment options should be investigated.

As such, there are limited treatment options for MG infections and monitoring of antimicrobial resistance to the most frequently used antibiotics is crucial to tailor treatment and testing guidelines. Here, we showed that the current surveillance method for MG AMR in Belgium is sub-optimal and should be adjusted. As in France, a prospective study annually or every other year, could provide a better estimation of the real magnitude of MG AMR.

Our study had several limitations. Firstly, the laboratories did not use the same testing strategy nor did they all have access to socio-demographic & behavioural data. The MG positivity ratios only provide rough estimates of prevalence as they are impacted by testing strategies applied in the different laboratories. The variation in positivity ratios reflect the sampling bias and they cannot be used as an accurate estimation of the MG prevalence in Belgium. Secondly, our sample size of MSM and MSW was limited. Finally, we did not have any information about the clinical response rate on treatment. As such, our resistance figures may be overestimated.

In conclusion, our study underscores the importance of correct surveillance of antimicrobial resistance of MG to adapt testing- and treatment guidelines. Macrolide resistance testing is expensive and currently not reimbursed in Belgium. We here show that this test may not be relevant among MSM, however, macrolide resistance testing for MG should be implemented as soon as possible in other populations to avoid further emergence of multidrug resistant MG.

## Data Availability

The data supporting the findings of this publication are retained at the Institute of Tropical Medicine, Antwerp and will not be made openly accessible due to ethical and privacy concerns. According to the ITM research data sharing policy, only fully anonymized data can be shared publicly. The data are de-identified (using a unique patient code) but not fully anonymized and it is not possible to fully anonymize them due to the longitudinal nature of the data. Data can however be made available after approval of a motivated and written request to the ITM at. The ITM data access committee will verify if the dataset is suitable for obtaining the study objective and assure that confidentiality and ethical requirements are in place.

## Acknowledgements

We would like to thank all laboratory personnel and physicians involved in the study. Moreover, we would like to specifically thank the personnel of the National Reference Centre for STIs.

## Contribution

DVDB, IDB, EP, AL, WVB, HS conceptualized the study. KK, IL, MR, IM, LF, MC, EVE, NM, GS, WV, BG, VM, YVdB, RC, SS, AL, MG, HP, EL, BL, MD, AH, VS provided samples for the study. HS performed antimicrobial testing. IDB performed the statistical analysis. IDB wrote the first draft of the manuscript. DVDB, HS, CK reviewed the first draft and all authors reviewed and provided comments to the final draft of the manuscript.

## References

[1] Baumann L, Cina M, Egli-Gany D, Goutaki M, Halbeisen FS, Lohrer G-RR, et al. Prevalence of Mycoplasma genitalium in different population groups: systematic review andmeta-analysis. Sex Transm Infect 2018;94:255–62. https://doi.org/10.1136/sextrans-2017-053384.

[2] Reyniers T, Nöstlinger C, Laga M, De Baetselier I, Crucitti T, Wouters K, et al. Choosing Between Daily and Event-Driven Pre-exposure Prophylaxis. JAIDS J Acquir Immune Defic Syndr 2018;79:186–94. https://doi.org/10.1097/QAI.0000000000001791.

[3] Coorevits L, Traen A, Bingé L, Descheemaeker P, Boelens J, Reynders M, et al. Macrolide resistance in Mycoplasma genitalium from female sex workers in Belgium. J Glob Antimicrob Resist 2018;12:149–52. https://doi.org/10.1016/j.jgar.2017.09.018.

[4] Hetem DJ, Kuizenga Wessel S, Bruisten SM, Braam JF, van Rooijen MS, Vergunst CE, et al. High prevalence and resistance rates of Mycoplasma genitalium among patients visiting two sexually transmitted infection clinics in the Netherlands. Int J STD AIDS 2021;32:837–44. https://doi.org/10.1177/0956462421999287.

[5] Manhart LE, Geisler WM, Bradshaw CS, Jensen JS, Martin DH. Weighing Potential Benefits and Harms of Mycoplasma genitalium Testing and Treatment Approaches. Emerg Infect Dis 2022;28:E1–11. https://doi.org/10.3201/EID2808.220094.

[6] Wood GE, Bradshaw CS, Manhart LE. Update in Epidemiology and Management of Mycoplasma genitalium Infections. Infect Dis Clin North Am 2023;37:311–33. https://doi.org/10.1016/j.idc.2023.02.009.

[7] Jensen JS, Cusini M, Gomberg M, Moi H, Wilson J, Unemo M. 2021 European guideline on the management of Mycoplasma genitalium infections. J Eur Acad Dermatology Venereol 2022. https://doi.org/10.1111/JDV.17972.

[8] Horner PJ, Blee K, Falk L, van der Meijden W, Moi H. 2016 European guideline on the management of non-gonococcal urethritis. Int J STD AIDS 2016;27:928–37. https://doi.org/10.1177/0956462416648585.

[9] Chow EPF, Walker S, Hocking JS, Bradshaw CS, Chen MY, Tabrizi SN, et al. A multicentre double-blind randomised controlled trial evaluating the efficacy of daily use of antibacterial mouthwash against oropharyngeal gonorrhoea among men who have sex with men: the OMEGA (Oral Mouthwash use to Eradicate GonorrhoeA) study protocol. BMC Infect Dis 2017;17:456. https://doi.org/10.1186/s12879-017-2541-3.

[10] De Baetselier I, Kenyon C, Vanden Berghe W, Smet H, Wouters K, Van den Bossche D, et al. An alarming high prevalence of resistance-associated mutations to macrolides and fluoroquinolones in Mycoplasma genitalium in Belgium: results from samples collected between 2015 and 2018. Sex Transm Infect 2021;97:297–303. https://doi.org/10.1136/sextrans-2020-054511.

[11] Pereyre S, Laurier-Nadalié C, Roy C Le, Guiraud J, Dolzy A, Hénin N, et al. Prevalence of macrolide and fluoroquinolone resistance-associated mutations in Mycoplasma genitalium in metropolitan and overseas France. Sex Transm Infect 2022:sextrans-2022-055466. https://doi.org/10.1136/SEXTRANS-2022-055466.

[12] Jensen JS. Protocol for the Detection of Mycoplasma genitalium by PCR from Clinical Specimens and Subsequent Detection of Macrolide Resistance-Mediating Mutations in Region V of the 23S rRNA Gene, Humana Press, Totowa, NJ; 2012, p. 129–39. https://doi.org/10.1007/978-1-61779-937-2_8.

[13] Shimada Y, Deguchi T, Nakane K, Masue T, Yasuda M, Yokoi S, et al. Emergence of clinical strains of Mycoplasma genitalium harbouring alterations in ParC associated with fluoroquinolone resistance. Int J Antimicrob Agents 2010;36:255–8. https://doi.org/10.1016/j.ijantimicag.2010.05.011.

[14] Sweeney EL, Whiley DM, Murray GL, Bradshaw CS. Mycoplasma genitalium: enhanced management using expanded resistance-guided treatment strategies. Sex Health 2022. https://doi.org/10.1071/SH22012.

[15] Aanpak van SOA’s door de eerste lijn n.d. https://www.soa.kce.be/en/treatment.html.

[16] De Baetselier I, Cuylaerts V, Smet H, Abdellati S, De Caluwe Y, Taïbi A, et al. Neisseria gonorrhoeae antimicrobial resistance surveillance report of Belgium – 2022. Antwerp: 2022.

[17] Piñeiro L, Idigoras P, Arrastia M, Manzanal A, Ansa I, Cilla G. Increases in the Macrolide Resistance of Mycoplasma genitalium and the Emergence of the A2058T Mutation in the 23S rRNA Gene: Clonal Spread? Antibiotics 2022;11. https://doi.org/10.3390/antibiotics11111492.

[18] Braam JF, Slotboom B, Van Marm S, Severs TT, Van Maarseveen NM, Van Zwet T, et al. High prevalence of the A2058T macrolide resistance-associated mutation in Mycoplasma genitalium strains from the Netherlands. J Antimicrob Chemother 2017;72:1529–30. https://doi.org/10.1093/jac/dkw584.

[19] Angela A, Raffaele DP, Federica R, Adriana M, Luigi S, Luigi R. Multi-year prevalence and macrolide resistance of Mycoplasma genitalium in clinical samples from a southern Italian hospital. Eur J Clin Microbiol Infect Dis 2021;40:893–5. https://doi.org/10.1007/S10096-020-04068-3/METRICS.

[20] Perry MD, Jones S, Bertram A, de Salazar A, Barrientos-Durán A, Schiettekatte G, et al. The prevalence of Mycoplasma genitalium (MG) and Trichomonas vaginalis (TV) at testing centers in Belgium, Germany, Spain, and the UK using the cobas TV/MG molecular assay. Eur J Clin Microbiol Infect Dis 2023;42:43–52. https://doi.org/10.1007/S10096-022-04521-5.

[21] Kenyon C, Vanbaelen T, Van C. Recent insights suggest the need for the STI field to embrace a more eco-social conceptual framework[]: A viewpoint. Int J STD AIDS 2022;published. https://doi.org/10.1177/09564624211064133.

[22] Murray GL, Bodiyabadu K, Vodstrcil LA, Machalek DA, Danielewski J, Plummer EL, et al. parC Variants in Mycoplasma genitalium: Trends over Time and Association with Moxifloxacin Failure. Antimicrob Agents Chemother 2022;66:1–5. https://doi.org/10.1128/aac.00278-22.

[23] Raccagni AR, Bruzzesi E, Spagnuolo V, Canetti D, Castagna A, Nozza S. ‘Multidrug-resistant Mycoplasma genitalium urethritis: successful eradication with sequential therapy. Sex Transm Infect 2023;99:77. https://doi.org/10.1136/SEXTRANS-2022-055678.

[24] Goodfellow JJ, Hughes S, Smith J, Jones R, Moore LSP, Rayment M. Novel use of oral chloramphenicol for treatment-resistant Mycoplasma genitalium. Sex Transm Infect 2023;99. https://doi.org/10.1136/SEXTRANS-2022-055621.

