## supplementary figure 1 for "Estimation of the real magnitude of antimicrobial resistance of *Mycoplasma genitalium* in Belgium by implementing a prospective surveillance programme"

**Supplementary material**

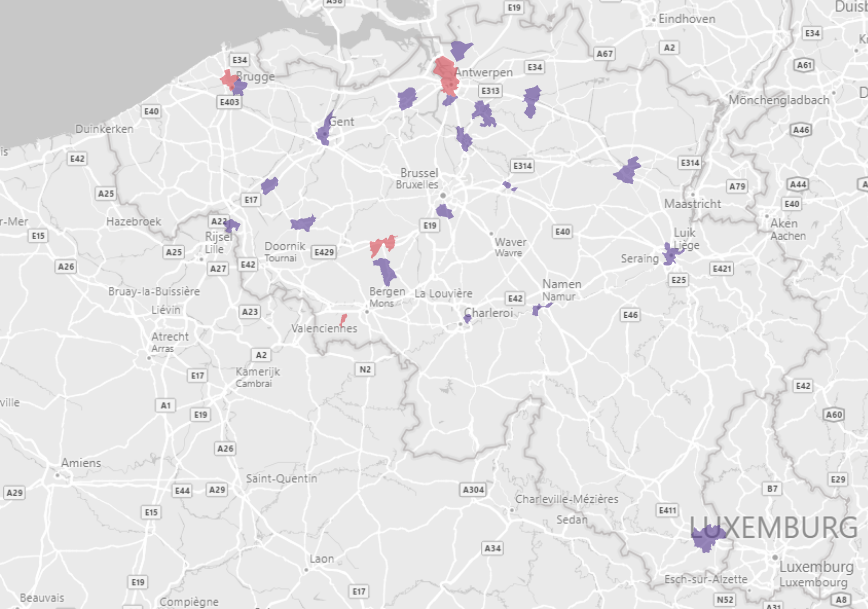

Supplementary Figure 1:Geographical location of laboratories participating in the prospective study. The red laboratories provided information about the prevalence of M. genitalium.

Supplementary table 1: Mycoplasma genitalium testing characteristics including positivity ratio of Belgian laboratories who participated in the study. The number of samples shipped for resistance testing is added in the last column

| Kind Of Laboratory | Geographical location | *M. genitalium* Molecular detection assay | Number of *M. genitalium* tests during Q1 2022 | Number of positive *M. genitalium* samples during Q1 2022 | *M. genitalium* Positivity ratio during Q1 2022 | Samples shipped for Resistance testing |
| --- | --- | --- | --- | --- | --- | --- |
| Private laboratory | Flanders | Abbott - Alinity | 1379 | 47 | 3.4% | 12 |
| Hospital laboratory | Flanders | Abbott - Alinity | 17 | 1 | 5.9% | 3 |
| Hospital laboratory | Flanders | Abbott - Alinity | 20 | 0 | 0.0% | 1 |
| Private laboratory | Wallonia | Abbott - Alinity | 5878 | 218 | 3.7% | 14 |
| Hospital laboratory | Wallonia | Diagenode MG/TV | 116 | 12 | 10.3% | 14 |
| Hospital laboratory | Flanders | Elite - STI PLUS MGB | 206 | 3 | 1.5% | 8 |
| Hospital laboratory | Flanders | Elite - STI PLUS MGB | 267 | 6 | 2.2% | 10 |
| Hospital laboratory | Wallonia | Elite - STI PLUS MGB | 14 | 0 | 0.0% | 1 |
| Hospital laboratory | Wallonia | Elite - STI PLUS MGB | 965 | 13 | 1.3% | 12 |
| Private laboratory | Flanders | Elite - STI PLUS MGB | 187 | 9 | 4.8% | 12 |
| Hospital laboratory | Flanders | Laboratory Developed Test (Taqman Array Card, Lifetech) | 1167 | 63 | 5.4% | 15 |
| Hospital laboratory | Wallonia | Mikrogen | 94 | 4 | 4.3% | 14 |
| Hospital laboratory | Wallonia | Neumodx - TV/MG | 43 | 6 | 14.0% | 3 |
| Hospital laboratory | Flanders | R-biopharm Ridagene STI Mycoplasma | 39 | 1 | 2.6% | 2 |
| Private Laboratory | Flanders | Roche - Cobas® TV/MG | 103 | 13 | 12.6% | 20 |
| Hospital laboratory | Wallonia | Seegene - Allplex CT/NG/MG/TV | 797 | 13 | 1.6% | 12 |
| Private laboratory | Wallonia | Seegene - Allplex CT/NG/MG/TV | 1034 | 69 | 6.7% | 12 |
| Hospital laboratory | Brussels | Seegene - Allplex CT/NG/MG/TV | 0 | 0 | Not Applicable | 12 |
| Hospital laboratory | Flanders | Seegene - Allplex CT/NG/MG/TV | 277 | 26 | 9.4% | 16 |
| Hospital laboratory | Flanders | Seegene - Allplex STI Essential Assay | 399 | 13 | 3.3% | 11 |
| Hospital laboratory | Flanders | Seegene - Allplex STI Essential Assay | 206 | 8 | 3.9% | 4 |
